# Using the Bayesian credible subgroups method to identify populations benefiting from treatment: An application to the Look AHEAD trial

**DOI:** 10.1101/2019.12.10.19014357

**Authors:** Anna Coonan, John Forbes, Patrick Schnell, Joel Smith

**Affiliations:** Graduate Entry Medical School, The University of Limerick, Limerick Ireland; Division of Biostatistics, College of Public Health, The Ohio State University, Columbus, OH, USA; Nuffield Department of Population Health, The University of Oxford, Oxford, UK

## Abstract

Traditionally, subgroup analyses are used to assess whether patient characteristics moderate treatment effectiveness with general disregard for issues of multiplicity. Using data from The Action for Health in Diabetes (Look AHEAD) trial in the United States, we aim to identify a subgroup where all of its types of members experience a treatment benefit defined as reducing the likelihood of a major cardiovascular event under an intensive lifestyle and weight-loss intervention. We apply the credible subgroups method to a Bayesian logistic model with a conservative prior that is sceptical of large treatment effect heterogeneity. The covariate profiles for which there is sufficient evidence of treatment benefit are, coarsely, middle-aged women, in poor subjective general health and with moderately to poorly controlled diabetes. There is at least 80% posterior probability that the conditional average treatment effect is positive for all covariate profiles fitting this description, which account for 0.5% of trial participants. Conversely, the covariate profiles that are likely to be associated with no benefit are middle aged and older men in excellent subjective general health, with well-controlled diabetes. These profiles apply to less than 2% of trial participants. More information is required to determine treatment benefit or no benefit for the remainder of the trial population.

## 1. Introduction

In general, healthcare providers aim to use information about a person to prevent, diagnose and treat disease, or to promote wellness. This relies on the ability to translate results from trials into clinical practice where the quantity of interest is usually the average treatment effect (ATE). The experimental approach to the estimation of average treatment effects involves random assignment of treatment followed by a comparison of the average outcomes of treated to nontreated individuals that serve as controls [1]. However, ATEs only provide a partial explanation of treatment effects particularly when they vary across experimental units.

In randomised controlled trials, average treatment effects tend to be generalised to a wider population and as such the results will be taken to apply equally to all patients in the trial. Participants, however, will vary significantly in baseline predictive and prognostic characteristics, and therefore each patient can report a unique effect with respect to a given treatment. This treatment effect heterogeneity can be masked by the calculation of average effects, leaving potential benefiters and non-benefiters of a given treatment unidentified. Producing results on population averages without recognizing the underlying heterogeneity could lead to welfare losses and increases in healthcare inefficiencies and costs [2, 3].

The standard approach for investigating treatment effect heterogeneity is to estimate conditional average treatment effects, or treatment effects among homogeneous groupings of experimental units. These homogenous subgroups are defined by baseline covariates. In order to assess how causal effects vary across subgroups, researchers predict conditional average treatment effects (CATEs) which are average effects conditioned on certain values of observed covariates for which treatment effectiveness varies [4]. Conditional average treatment effects are particularly useful when the researcher wants to explore the heterogeneity of treatment effects in various subgroups defined by a single covariate.

Estimating heterogeneous treatment effects, or systematically different outcomes associated with treatment due to moderating patient characteristics, is typically relegated to subgroup analyses that rely on simple treatment-covariate interaction terms [5]. In clinical trials, however, researchers typically aren’t concerned with just one subgroup but rather subgroups across a series of standard covariates such as age, ethnicity, and sex. Multiple interaction tests can be misleading as they are often underpowered, suffer from multiplicity errors and do not allow for simultaneous inference [6, 7]. Moreover, subgroups tend to be prespecified to avoid perceptions of data-mining or overfitting the data and therefore restrict the ability of the researcher to test new unforeseen hypotheses [8].

There is growing recognition in the field of health economics that more nuanced estimates of patient-specific heterogeneous treatment effects can be estimated and conditioned on a variety of observable and unobservable risk factors. Some methods are derived from the computer science discipline such as machine learning methods where the focus is on predicting patient-specific treatment effects [9-11]. Other methods build on a Bayesian framework where the parameter of interest is unknown but the model allows the researcher to vary the degree of expected treatment effect heterogeneity through *a priori* selection [6, 12, 13]. All of these methods provide insight into the reliability of incorporating heterogeneity in treatment response.

Our work builds on a Bayesian framework to classify personalised treatment effects (PTEs), or average outcomes under treatment and control for every covariate point. The PTEs are used to classify covariate points into credible subgroups that allow for inferences about the types of patients expected to benefit or not benefit from an intervention, while accounting for the multiplicity arising from testing a hypothesis at every covariate point. This study is the first major application of the Bayesian credible subgroups method to such a large clinical dataset. Previous research and our own are discussed in the context of a large multicentre randomised controlled trial called the Action for Health in Diabetes (Look AHEAD). In the Look AHEAD trial, insignificant treatment effects for the whole sample and all prespecified subgroups were reported, most likely resulting from latent patient heterogeneity [14].

In 2001 the Look AHEAD trial set out to model the long-term effects of an intensive lifestyle intervention (ILI), one specifically aimed at weight loss on cardiovascular morbidity and mortality among patients with type 2 diabetes who were also overweight or obese [14, 15]. The ILI included the following: individual and group counselling sessions on diabetes control, aids for calorie management with the option of meal replacement supplements, and supports to increase physical activity. Patients in the control group received diabetes support and education (DSE) counselling only. Prior to the Look AHEAD trial, the effects of lifestyle interventions on cardiovascular morbidity and mortality were mixed at best. Some studies reported no cardiovascular effect and others reported minimal benefits after two decades of follow-up [16-19]. The effects of weight loss on reduced cardiovascular risk factors in the short-run were well-documented, but the Look AHEAD trial set out to identify if significant effects translated to reductions in a primary composite CVD endpoint over a long-term follow-up. In September 2012 the Look AHEAD trial prematurely ended after a futility analysis concluded the weight loss intervention did not reduce the rate of primary cardiovascular events [14]. The median follow-up was 9.6 years. Data are available upon request from the National Institute of Diabetes and Digestive and Kidney Diseases (NIDDK). All participants in Look AHEAD provided written consent to allow their information to be used in future research.

A possible explanation for a lack of statistical significance in Look AHEAD was the presence of differing effects for different individuals or patient heterogeneity [14]. Secondary analyses were conducted to see if general health benefits resulted from the lifestyle intervention. The documented health benefits range from weight loss, increased physical fitness, better diabetes control, and reduced urinary incontinence [20-22]. A small number of exploratory studies have examined the original CVD endpoint finding positive treatment effects for subgroups defined by subjective general health and diabetes control [23] and percentage weight loss in the first twelve months [20]. Using different methods, these were all attempts to model treatment heterogeneity in Look AHEAD.

Behavioural responses and treatment uptake are notoriously heterogeneous with varying degrees of weight loss being reported in the first year [24]. In the instance of the Look AHEAD trial it is likely that patient heterogeneity was able to moderate treatment effectiveness at two entry points: the effect of the intervention on behaviour and uptake, and the effect of behaviour and weight loss on the primary CVD outcome [25]. In Baum et.al. [23] a machine learning algorithm based on decision-trees was used to identify two baseline patient characteristics believed to be instigators behind the trial-wide outcome neutral effects. One covariate was HbA_1c,_ or glycated haemoglobin, which is a measure of diabetes control. The second instigator of treatment effect heterogeneity was a subjective general health indicator derived from the SF-36 questionnaire. These covariates were determined to be the most important variables for defining the range of patient heterogeneity. Defined by HbA_1c_ and subjective general health, two subgroups with positive average treatment effects were identified. However, the HbA_1c_ and subjective general health (SF-36) compositions within these two subgroups were contradictory. One subgroup reported a significant absolute risk reduction defined by the baseline covariates excellent subjective general health (SF-36 ≥ 48) and well-controlled diabetes (HbA_1c_ < 6.8%). After participants with an expected adverse treatment response were removed from the analysis, a second subgroup with significant risk reductions was defined by the baseline covariates HbA_1c_ ≥ 6.8% or both HbA_1c_ < 6.8% and SF-36 ≥ 48. These results led to conclusions stating 85% of Look AHEAD participants averted a cardiovascular event which would be a large overestimation of treatment benefit compared to previous Look AHEAD studies [14, 20, 25, 26]. The mixed composition of these subgroups support previous criticisms stating too much emphasis is placed on the coefficients produced my machine learning algorithms and that they are not designed to collate patients into subgroups. Rather, machine learning methods are better suited for examining unit-level heterogeneity [27].

Machine learning methods are useful for detecting and analysing heterogeneity, but they do not directly address the question of primary interest to the current study: specifically whom does treatment benefit? Previously-applied machine learning methods do not adjust for multiplicity or provide simultaneous inferences, defined as a situation where all types of individuals within a subpopulation satisfy a common statement [6]. The most relevant statement here is that all covariate points in the subgroup correspond to a positive PTE. We use a Bayesian credible subgroups method for simultaneous inference that through careful choice of priors on treatment-covariate interactions, allows us to tune our *a priori* belief in the strength of treatment effect heterogeneity. Our model includes a conservative prior that is sceptical of large treatment effect heterogeneity but still allows for strong evidence from the posterior distribution to overwhelm the prior information.

The inferential goal of this analysis is different from previous secondary Look AHEAD studies that confirm the overall treatment effect is neutral and any subsequently identified subgroups are defined by estimated average treatment effects. These estimates continue to apply assumptions of treatment effect homogeneity within all levels of selected covariates. Instead, we aim to capitalise on inherent patient heterogeneity in order to identify the subgroup consisting of only types of patients expected to benefit, not a subgroup of any types patients that together yield a positive average effect. We explore the mechanisms behind the statistical insignificance of the lifestyle intervention. This is done by examining the possibility of a treatment-benefiting subgroup defined by the baseline covariates sex, age, blood glucose percentage (HbA_1c_) and a measure of subjective general health (SF-36).

This article is structured as follows: In Section 2 we present the methodology and selection of priors. Section 3 describes the data from the Look AHEAD and defines the descriptive statistics. Section 4 presents the results from the credible subgroups analysis with varying credible levels. Section 5 concludes with a discussion and suggestions for future research.

## 2. Methodology

Data provided by the NIDDK repository was fully anonymised before we accessed them. Our research received ethical approval from University of Limerick application number 2017_12_19_EHS. All participants in Look AHEAD provided written consent to allow their information to be used in future research.

This analysis uses the Bayesian credible subgroups method to construct two bounding subgroups for a benefiting subgroup B that is unknown. The benefiting subgroup is defined to consist of all covariate profiles with a conditional average treatment effect above a threshold set to zero (*δ* = 0), intended to capture all positive treatment effects regardless of magnitude. Inferences about the subgroup B are made by constructing bounding subgroups, D and S, similar to the procedure for making inferences about a scalar quantity by constructing interval estimates. The bounding subgroups are constructed with the goal that one, the exclusive credible subgroup (D), contains only covariate profiles with a positive treatment effect. The other, the inclusive credible subgroup (S), is intended to contain all covariate profiles with a positive treatment effect. In the construction of these bounds, treatment effects are estimated and tested at all points throughout the covariate space, thereby introducing multiple testing issues. However, by estimating a subgroup where all of its members satisfy some criteria, such as having a treatment effect above some threshold of significance, this statement of simultaneity accounts for multiplicity due to performing a hypothesis test at each covariate point. The result is a decreased risk of conducting a family wise type I error.

The method of credible subgroups is a two-step regression classification method. Parameters of a linear or non-linear regression model are estimated, and a simultaneous credible band of some level around the regression surface is constructed. Covariate points with a lower bound above some threshold form the exclusive credible subgroup (D), and those where the upper bound is greater than the threshold become the inclusive credible subgroup (S). As an example of the first step, estimation of a standard linear model includes predictive and prognostic covariates generally described as follows:

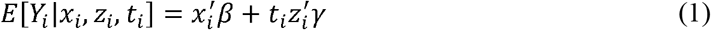

Y is the response, *x* and *z* are vectors of prognostic and predictive covariates where predictive covariates are those interacted with treatment allocation. *β* and *γ* are the effect parameters and *t* ∈ (0,1) the treatment allocation. Additionally, the model includes a conservative prior on *y* that represents the belief that treatment-covariate interactions are unlikely by shrinking coefficients towards zero, but that still allows for strong evidence of treatment effect heterogeneity from the posterior distribution to overrule this. The second step, classification, comes from the posterior distribution of patients’ personalised treatment effects (PTEs). These are used to classify all combinations of covariates in the covariate space according to a credible subgroup pair: the exclusive credible subgroup (D) and the inclusive credible subgroup (S). PTEs provide a clinically useful interpretation of the expected difference in outcomes for patients that share the same baseline characteristics.

In our analysis the covariate space is comprised of age, sex, subjective general health measured by the SF-36 general health questionnaire, and diabetes control measured a glycated haemoglobin percentage (HbA_1c_). All of the covariates included in this paper are standard medical and demographic variables included in studies examining behavioural treatments for type 2 diabetes and the confounding effects of obesity [14, 17, 18, 20, 23, 28]. The model of the personalised treatment effect is the difference in the expectation of the outcome (*y*) conditional on the predictive covariates (*z*), as follows:

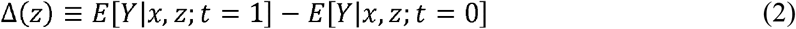

From the posterior distribution of personalised treatment effects the covariate space is divided into three parts based on a decision rule used to define the credible subgroup pair (D,S). This is comprised of the covariate profiles with lower bounds for treatment effects above some threshold (*δ* > 0) that define subgroup D, those in a complement subgroup S^c^ with upper bounds for treatment effects at most equal to the threshold (*δ* = 0) and an uncertainty region S\D (S remove D) where more information is required for classification. The trichotomy of the covariate space is illustrated in Fig 1. The defining property of the credible subgroups is as follows:

**Fig_1.**
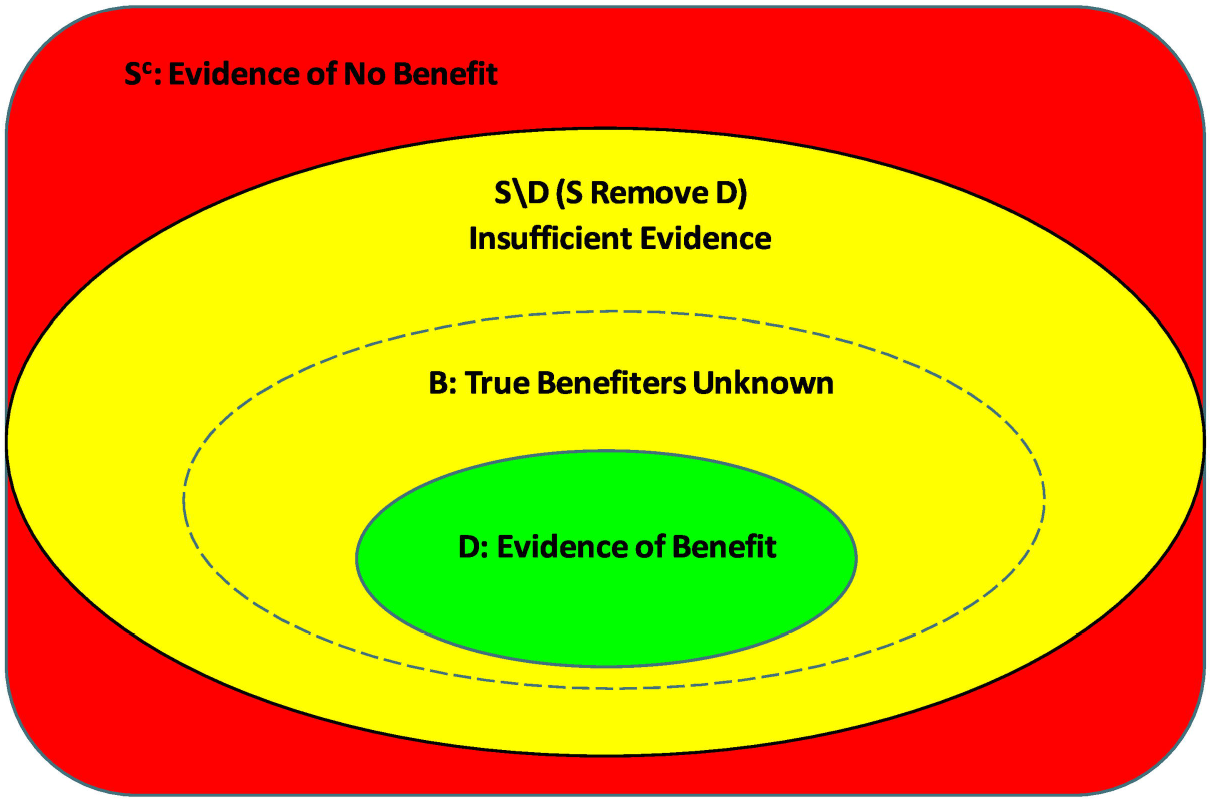
Trichotomy of Predictive Covariate Space. Visualisation of the trichotomy of the covariate space based on the credible subgroup pair (D,S) relative to the true benefiting subgroup (B), adapted from Schnell et.al. [6]

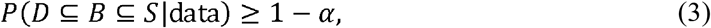

where D and S are subsets in the covariate space with a bound on the posterior probability that these two sets bracket the benefiting subset B. Formally, D is the exclusive credible subgroup because the posterior probability that D contains only z for which Δ(*z*) > *δ*, and simultaneously, S is the inclusive subgroup that contains all z such that Δ(*z*) > *δ*, is at least 1 − *α*. The credible level of the subgroup pair (1 − *α*) is also an approximate lower bound on the frequentist probability that *D* ⊆ *B* ⊆ *S*. In this study, D is the reported estimate with posterior probability 1 − *α* that the reported subset characterises exclusively covariate combinations with an expected treatment effect above the threshold (*δ* = 0).

The objective of this analysis is to identify a credible subgroup pair such that, at a credible level of at least 80%, D represents a subgroup that contains *only* types of patients with a positive treatment effect and S represents a subgroup that contains *all* types of patients with a positive treatment effect. Defining an exclusive credible subgroup (D) where all members have a treatment effect above some threshold satisfies properties of simultaneous inference and adjusts for multiplicity [6]. The credible subgroups method allows for inferences that state it is with high probability that the personalised treatment effect (Δ(z)) is positive at all covariate points (z) in the exclusive credible subgroup (D).

The primary endpoint in Look AHEAD is a binary composite variable measuring the presence or absence of at least one CVD outcome. We fit a Bayesian logistic regression model with the endpoint likelihood *Y*_*i*_|*η*_*i*_ ∼ *Bernoulli* (*logit*^−1^, *η*_*i*_) where 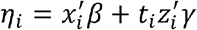. We use the vague prior *β*∼ *Normal* (0, 10^3^) for prognostic effects and a conservative prior *λ*∼ *Normal* (0,1) for predictive effects. In other words, the prognostic effect variances are set to 1000 and those corresponding to predictive effect variances set to 1. The benefiting subgroup is one for which the ILI treatment is superior to the diabetes support and education (DSE) control. This is in line with current clinical recommendations and the belief that physical activity and controlled caloric intake is beneficial for patients with type 2 diabetes who may be overweight or obese. We set the threshold of a log odds ratio of primary CVD occurrence to −*δ*_1_ > 0. The sign is switched for *δ* because we want reductions in risk of the primary CVD endpoint. Due to the insignificant results reported in the original trial, we wish to capture treatment benefits of any magnitude in order to avoid automatically eliminating covariate profiles that may not achieve a pre-defined level of interest. The model is fit using the NIMBLE package [29] in the R software [30] with 10,000 Gibbs sampler iterations after 1000 burn-in iterations. Convergence appears to be mixing well and near-immediate from autocorrelation plots for Markov Chains. Our analysis is conducted using only complete cases eliminating 8 observations which renders a sample size of 4893.

## 3. Data

Look AHEAD was a multicentre randomised controlled trial that tested the effect of an intensive lifestyle intervention focused on diet and exercise on long-term cardiovascular disease outcomes among patients with type 2 diabetes who were overweight or obese [14, 15]. Between August 2001 and April 2004 the trial recruited 5145 participants to undergo randomisation. 2570 were assigned to the intensive lifestyle intervention (ILI) group and 2575 were assigned to the diabetes support and education control (DSE) group. The intervention was modelled on group behaviour programs tailored for obese and overweight patients through individual and group counselling sessions. These sessions continued for an initial 6 months and decreased in frequency throughout a 12-month follow-up. Intervention design was tailored for a weight-loss goal of losing 10% initial body weight in the first year and a total weight-loss of 7% (or more) initial body weight by trial’s end. This would be achieved by limiting caloric intake with the option of meal replacement supplements and increasing physical fitness through weekly exercise regimes. The diabetes support and education (DSE) control group provided nutrition and exercise training for a period of four years after randomisation.

The primary outcome was a composite indicator of CVD morbidity and mortality consisting of CVD death, non-fatal myocardial infarction (MI), non-fatal stroke, and hospitalization for angina [14]. The number of CVD events was a concern throughout the trial resulting in the fourth CVD indicator, hospitalisation from angina, being added to the primary outcome two years into the trial [31]. The event rate of the CVD endpoint ended up being less than half the projected rate in the DSE control group, resulting in a very low overall rate of events [14]. Insignificant treatment effects were reported for secondary CVD composite measures and for all prespecified subgroups including age, sex and ethnicity.

Table 1 presents the descriptive statistics for patients in the treated and control groups of the remaining 4901 trial participants after 244 were omitted early in the trial for consent reasons. The characteristics of the patients in the two groups were similar at baseline. The ILI treatment group was on average 58.8 years old, had a BMI of 35.9, weighed 100.9kg and was 41% men. The DSE control group was on average 59.1 years old, had a BMI of 36.0, weighed 101.2kg and was 41% men. For the entire sample 75% of patients had moderately or poorly controlled diabetes defined as having an HbA_1c_ percentage greater than or equal to 6.5.

**Table 1.** Characteristics of Patients at Baseline

|  | Intensive Lifestyle Intervention (ILI)<br>(n = 2448) | Diabetes Support and Education (DSE)<br>(n = 2453) | Entire Trial Sample<br>(n = 4901) |
| --- | --- | --- | --- |
| Age (years) | 58.8 (6.7) | 59.1 (6.8) | 58.9 (6.8) |
| Male | 1014 (41%) | 1016 (41%) | 2030 (41%) |
| History of cardiovascular disease | 355 (15%) | 335 (14%) | 690 (14%) |
| Diabetes duration (years) | 6.7 (6.5) | 6.7 (6.3) | 6.7 (6.4) |
| Insulin use | 362 (15%) | 385 (16%) | 747 (15%) |
| Weight (kg) | 100.9 (19.6) | 101.2 (18.8) | 101.1 (19.2) |
| BMI (kg/m<sup>2</sup>) | 35.9 (6.0) | 36.0 (5.7) | 36.0 (5.9) |
| HbA<sub>1c</sub> (%) | 7.2 (1.1) | 7.3 (1.2) | 7.3 (1.2) |
| HbA<sub>1c</sub> (>=6.5%) | 1798 (73%) | 1867 (76%) | 3665 (75%) |
| Systolic blood pressure (mm Hg) | 128.5 (17.2) | 129.8 (17.1) | 129.1 (17.2) |
| SF-36 General Health (GH) Questionnaire | 47.2 (9.1) | 47.3 (8.7) | 47.2 (8.9) |

Data are in mean (SD) or n (%)

## 4. Results

The credible subgroups method is used to divide the covariate space into a trichotomy which will identify the profiles of patients expected to benefit from treatment, those expected not to benefit from treatment and those for whom more information is needed. The Look AHEAD trial is used in our analysis and includes an experimental intensive lifestyle intervention (ILI) versus a diabetes support and education (DSE) control. The covariates are believed to be both prognostic and predictive and are defined by baseline measurements for subjective general health measured by the SF-36 questionnaire, age, sex and diabetes control (HbA_1c_). The endpoint of interest is a binary composite indicator measuring the presence of four possible macro cardiovascular events.

The first stage of the credible subgroups method is the estimation of a Bayesian logistic regression with vague priors on prognostic effects and conservative priors on predictive effects. From this stage our objective is to discuss treatment-covariate interactions at the nominal credible level of 95% for which standard subgroup analyses were not powered to detect. The model is fit with 10,000 Gibbs sampler iterations after 1000 burn-in iterations. The posterior mean, standard deviations and credible bounds of effect parameters are reported in Table 2. Significant effects are reported for the treatment-control interaction and individual covariates general health and HbA_1c_. These results are not adjusted for multiplicity and are considered incomplete because a lack of significance on the age and sex interaction terms does not imply a homogeneous treatment effect across all levels of these covariates. The estimates in Table 2 do not identify the covariate profiles of patients explicitly expected to benefit and therefore do not directly address our research question. The interpretation that we wish to avoid is that the treatment only interacts with general health and HbA_1c_ and that any potential treatment effect exists across all levels of the other covariates.

**Table 2.** Posterior Summaries of Bayesian Logistic Parameters

|  | **Posterior**<br>**Mean** | **Posterior Standard**<br>**Deviation** | **95% Credible**<br>**Bounds** |
| --- | --- | --- | --- |
| Intercept | -2.41* | 0.14 | -2.18 to -2.63 |
| General Health | -0.07 | 0.06 | 0.03 to -0.17 |
| Age | 0.43* | 0.06 | 0.34 to 0.53 |
| Sex | 0.82* | 0.11 | 0.64 to 0.99 |
| HbA<sub>1c</sub> | 0.39* | 0.13 | 0.17 to 0.61 |
| Treatment | -0.22 | 0.18 | -0.52 to 0.07 |
| General Health ×<br>Treatment | -0.19* | 0.08 | -0.05 to -0.32 |
| Age × Treatment | -0.01 | 0.08 | -0.15 to 0.13 |
| Sex × Treatment | -0.07 | 0.16 | -0.33 to 0.18 |
| HbA<sub>1c</sub> × Treatment | 0.32* | 0.18 | 0.02 to 0.61 |

Continuous covariates are standardized. Estimates identified as significant according to a 95% credible level are denoted by \*.

Fig 2 presents the covariate space which constitutes the exclusive credible subgroup D at the 80% credible level. Credible subgroup membership was determined using the credsubs package [32]. This analysis uses a Bayesian logistic model to estimate posterior mean treatment effects from endpoint likelihoods that follow a Bernoulli distribution. Ellipsoidal contours of the standard errors are centred on a majority of the data points which induce the curved boundaries of the credible subgroup [6]. The covariate profiles classified to the exclusive credible subgroup D are middle-aged women in poor subjective general health with moderately to poorly controlled diabetes. There is at least 80% posterior probability that the conditional average treatment effect is beneficial (*δ* < 0) for all types of patients with covariate points in D, accounting for multiplicity due to testing a hypothesis for each covariate point, and thus promoting the lifestyle intervention as an appropriate treatment for this subgroup. Conversely, the S-complement subgroup identifies the covariate profiles of those who may not benefit, represented by either a null or adverse treatment effect (*δ* ≥ 0). The covariate profiles of patients in this region are middle-aged and older men, in excellent subjective general health with well-controlled diabetes. Therefore, the lifestyle intervention is not to be considered an appropriate treatment for members of this subgroup. A figure of similar threshold and credible level with a vague prior on predictive and prognostic effects is included in the Supplement S_Fig1. This figure supports the conclusions presented in Table 2.

**Fig 2.**
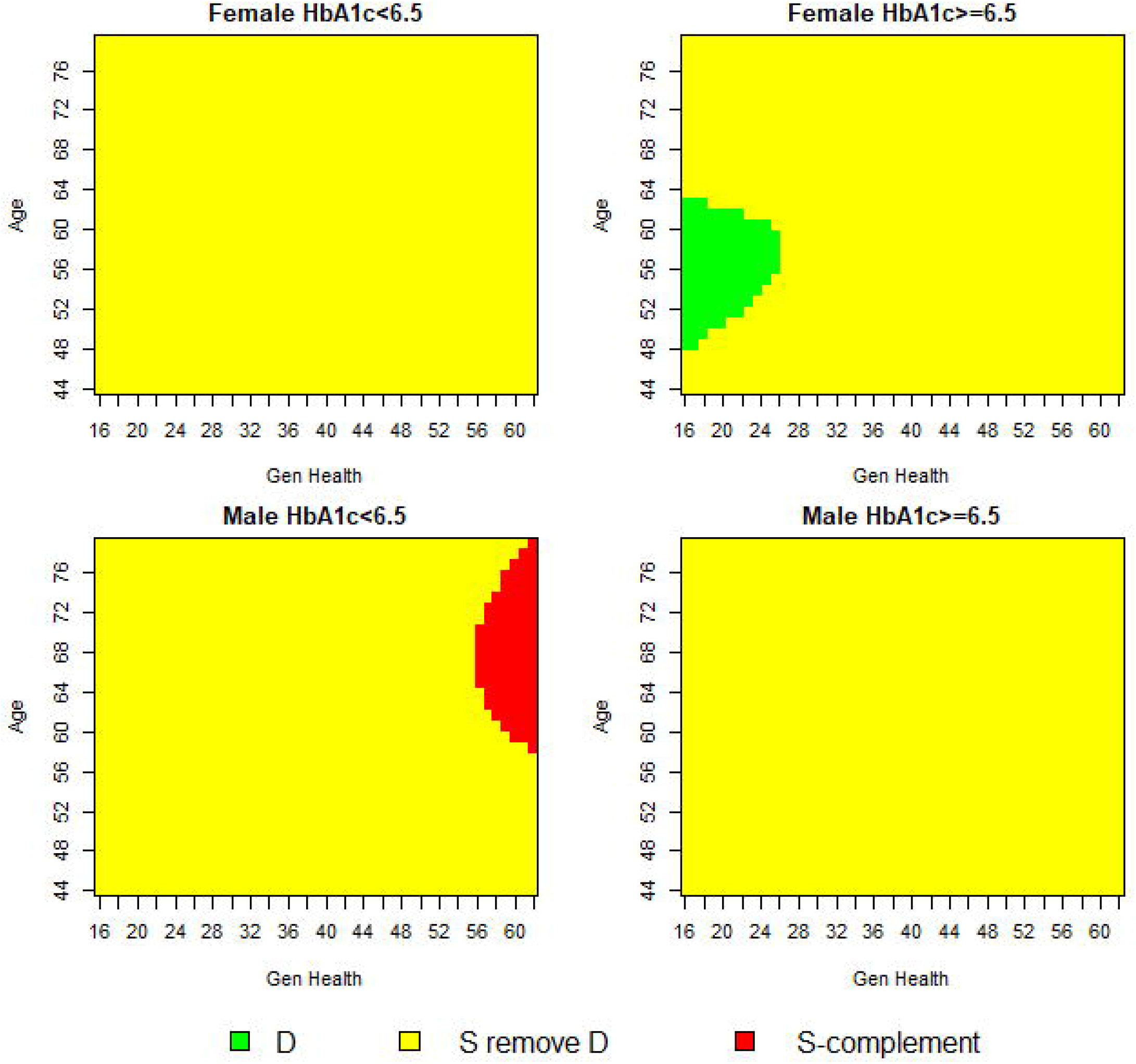
Trichotomy of Baseline Covariates at 80% Credible Level.

The funnel plot in Fig 3 shows the predicted treatment effects across the entire covariate space and their coloured classification according to the credible subgroups pair (D,S). A majority of covariate space is classified as the S \ D region where more information is required to classify points as being members of the exclusive credible subgroup. The funnel plot displays the proportion of covariates that are members of the S-complement subgroup and have a treatment effect at most equal to zero. This graph identifies the minority of covariate profiles with sufficient evidence of an expected treatment benefit represented by the green area. The minority of points classified to the exclusive credible subgroup supports the idea that the heterogeneity of patients with these covariate profiles could have been lost in the original trial-wide average estimates. When applying the trichotomy of the covariate space to 4893 observations in the Look AHEAD trial, there is sufficient evidence of benefit for less than 0.5% of the Look AHEAD population. Likewise, there is sufficient evidence of an expected null or adverse effect for less than 2% of the trial population.

**Fig 3.**
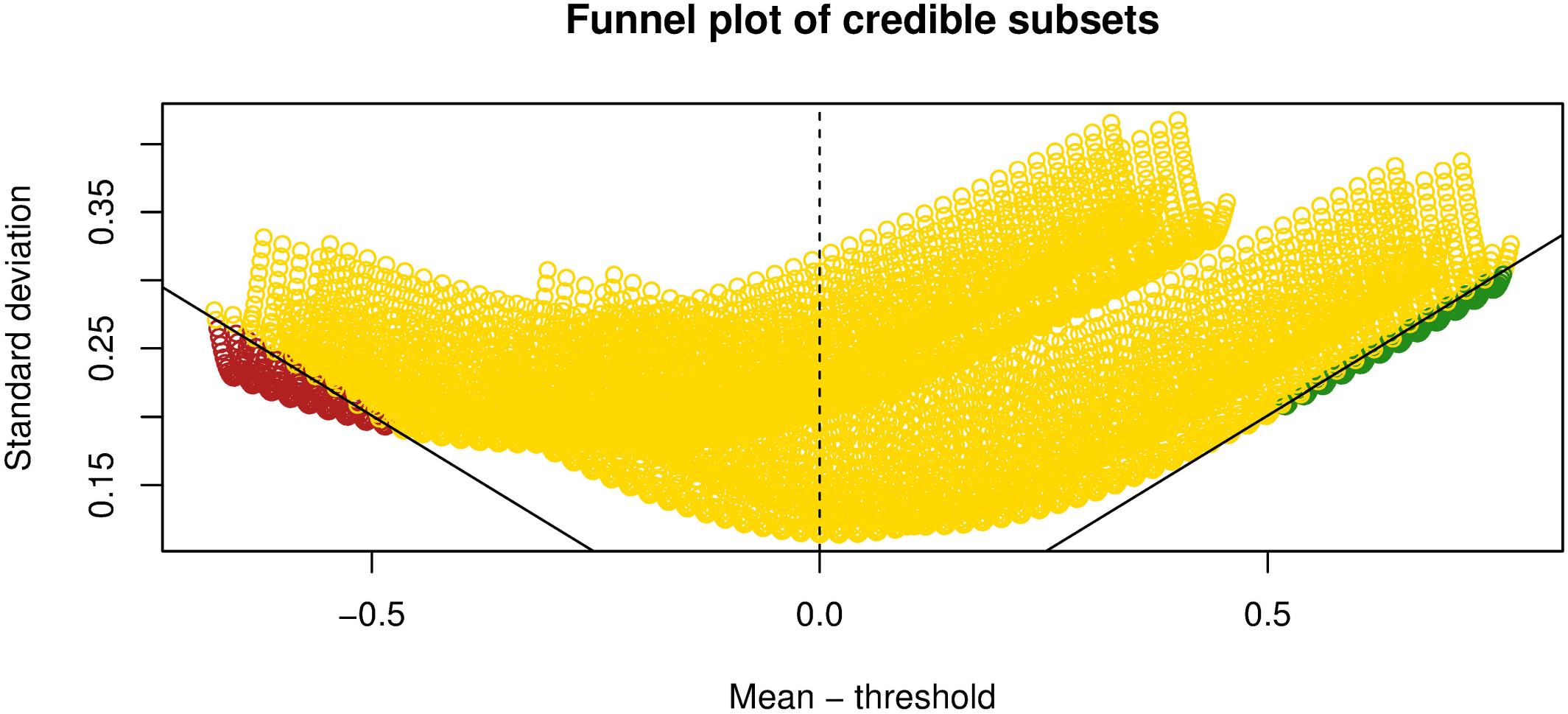
Exclusive (Green) and Inclusive (Yellow) Credible Subsets at 80% Credible Level.

Fig 4 shows the contours of the maximum credible level that allows assignment of a covariate point to the exclusive credible subgroup. Contours with negative values indicate assignment to the complement of the inclusive credible subgroup at the credible level of the modulus. The 80% contour aligns with the previously identified boundary of the exclusive credible subgroup shown in Fig 2. This figure displays the trade-off that exists in statistical reliability versus restrictiveness of subgroup composition. As the maximum credible level is relaxed, a greater portion of the covariate space is classified in either the exclusive credible subgroup or the complement of the inclusive credible subgroup.

**Fig 4.**
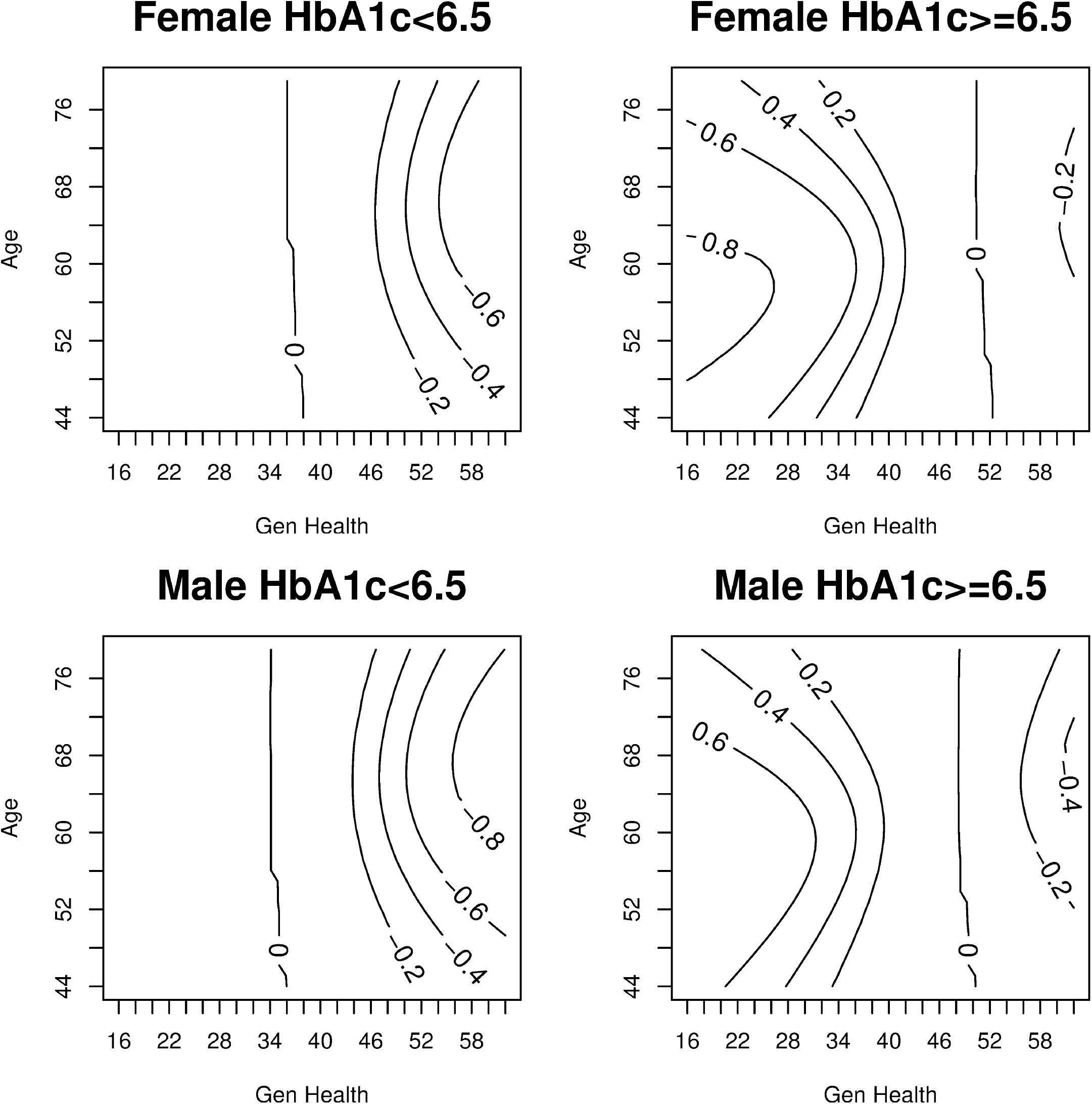
Maximum Credible Level Contours for Exclusive Credible Subgroup Inclusion.

In Fig 5 we set the treatment effect threshold to zero but relax the credible levels from 80% to 50%. A figure illustrating the trichotomy of the covariate space at a 65% credible level is included in the Supplement S_Fig2. As expected, when the credible level is relaxed, the proportion of covariate points allocated to the exclusive credible subgroup (D) and S-complement subgroup (S^c^) increase. As the credible level is relaxed, the covariate composition of the exclusive credible subgroup widens to include both men and women for a wider range of ages. The two most important covariates for differentiating patient benefiters from non-benefiters are having a low HbA_1c_ percentage and having good subjective general health. The similarity between these two covariates is to be expected as good control over one’s diabetes, which is a physical health indicator, could translate to patients’ positive subjective evaluations of their own health. This conclusion supports previous research that has found HbA_1c_ and subjective general health (SF-36) are two of the most important covariates for determining treatment effect heterogeneity in the Look AHEAD trial [23]. The HbA_1c_ composition of the exclusive and inclusive credible subgroups is largely consistent across sex, age and general health covariates.

**Fig 5.**
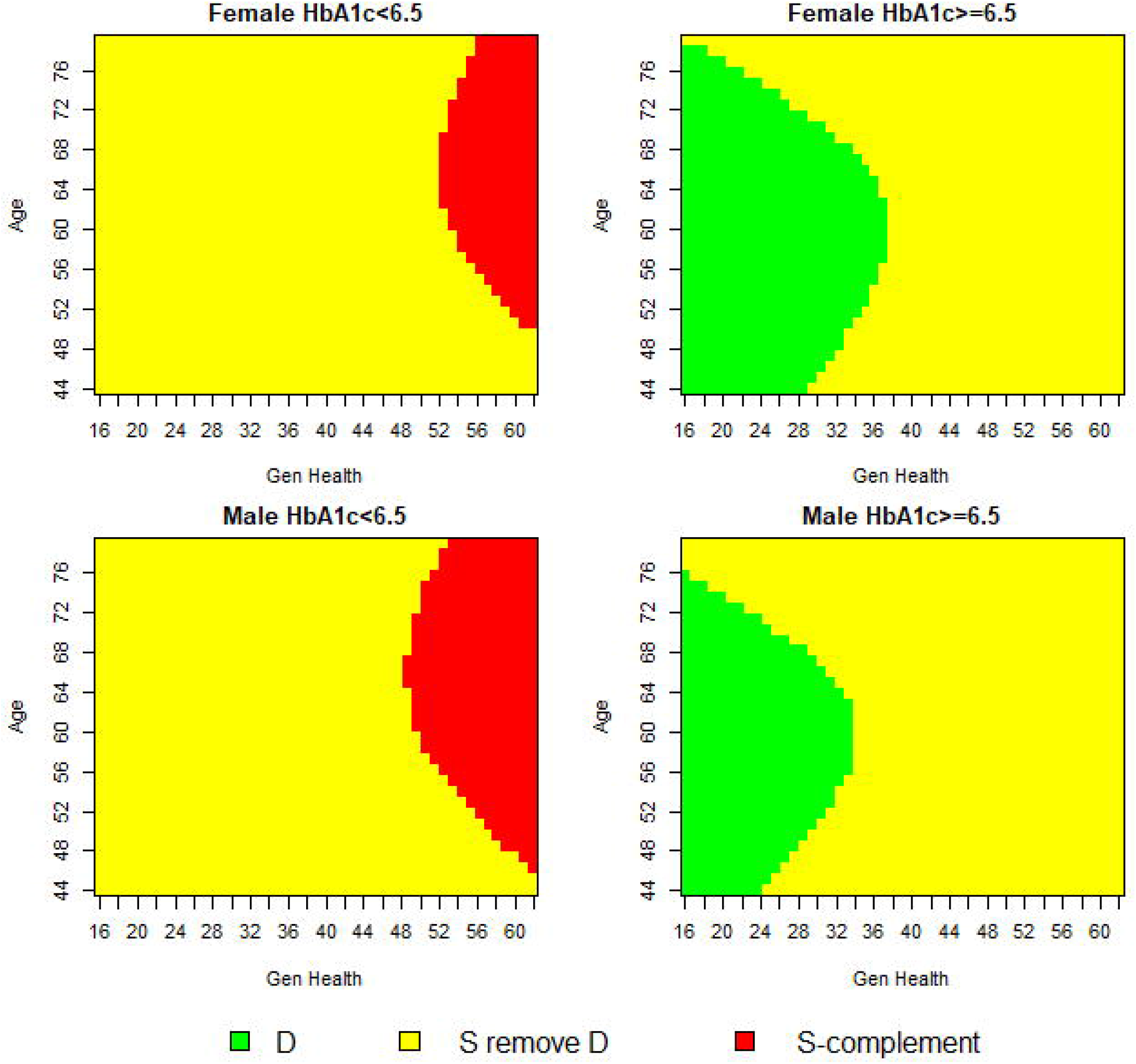
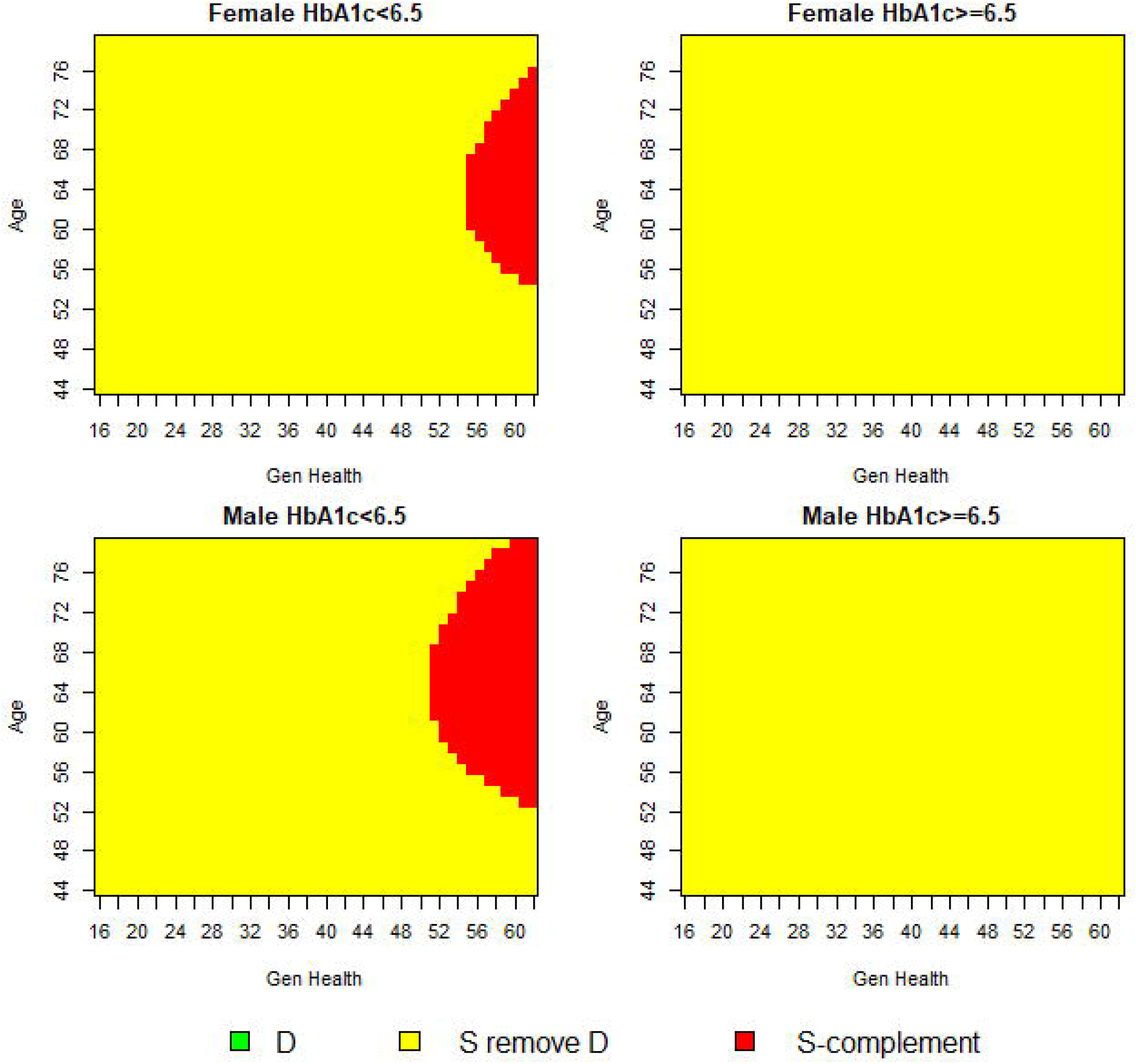
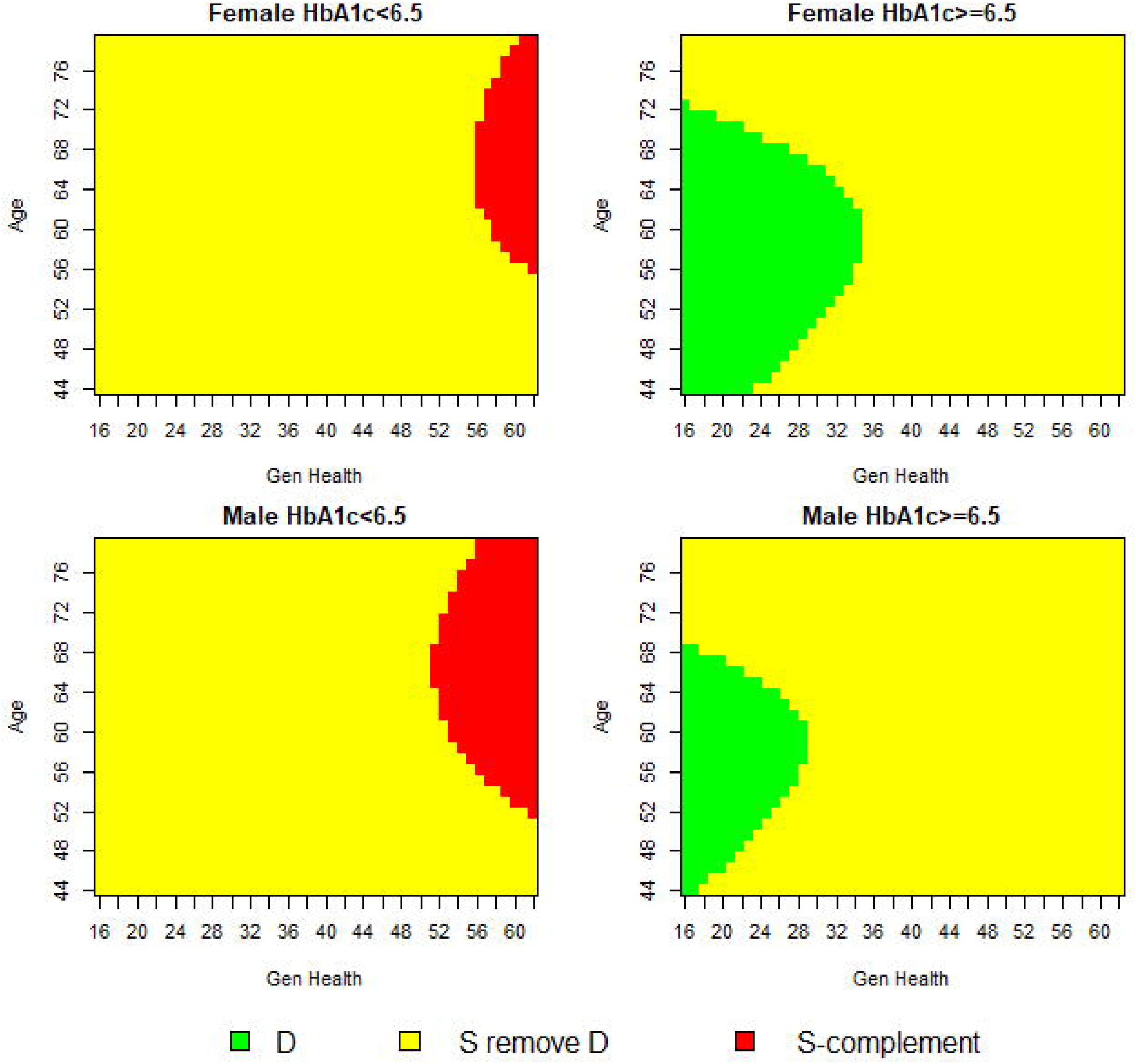
Trichotomy of Baseline Covariates at 50% Credible Level.

At the 80% credible level, the application of the covariate profiles of the exclusive credible subgroup, suggests evidence of treatment benefit for less than 0.5% of the Look AHEAD population. Applying the covariates that constitute the S-complement subgroup suggests evidence of a null or adverse treatment effect for 2% of the Look AHEAD population. At the more lenient 50% credible level, the application of the covariate profiles of the exclusive credible subgroup would render a subpopulation of less than 10% of the Look AHEAD trial. While the true benefiting population may be larger, the expansive inconclusive region across two credible levels does not provide supporting evidence of this. Our results across all credible levels differ from previous controversial findings that suggested as much as 85% of the Look AHEAD population benefitted from the ILI based on positive average estimates [23].

## 5. Discussion

In this analysis of the Look AHEAD trial, we identified a subgroup that benefited from an intense behavioural lifestyle intervention in terms of reduced cardiovascular risk. The aim of the trial was to model the effects of a lifestyle intervention that focused on weight-loss and physical activity on the long-term cardiovascular outcomes of patients with type 2 diabetes who were overweight or obese. After the original trial reported no between group differences in the occurrence of the primary CVD endpoint for patients randomised to the treatment or control, commentators suggested this was the result of latent patient heterogeneity. Recent developments in treatment effect estimation call for a move away from *ad hoc* subgroup analyses and encourage the development of new methodologies designed to capture the richness that patient heterogeneity provides. By fitting a Bayesian logistic regression model, we were able to include prior information of the degree of treatment effect heterogeneity and still allow for strong evidence from the posterior distribution to overrule any assumptions. Using the credible subgroups method, we classified covariate profiles according to a credible subgroup pair that allowed for the identification of a subgroup in which all types of patients exhibit a positive treatment effect. This subgroup was coarsely comprised of the following covariate profiles: middle-aged women in poor subjective general health with moderately to poorly controlled diabetes (HbA_1c_ ≥ 6.5%). There exists at least 80% posterior probability that the treatment effect is positive for patients with these covariate profiles located within the exclusive credible subgroup (D), accounting for multiplicity due to testing a hypothesis for each covariate point. A second subgroup was identified as the S-complement where its members exhibited a null or adverse treatment effect. The covariate profiles of this group were middle-aged and older men, in excellent subjective general health with well-controlled diabetes (HbA_1c_ < 6.5%). Applying the covariate profiles of the exclusive credible subgroup to the Look AHEAD participants rendered an eligible subpopulation of less than 0.5%. Members of this subpopulation would be expected to experience a positive treatment effect.

The covariate composition of the exclusive credible subgroup is consistent with studies from similar trials that reported women and those in poor health experienced the greatest reduction in CVD mortality after participating in a weight loss intervention [17, 33, 34]. Additionally, the identification of a S-complement group supports previous findings that a selection of patients can have null or adverse cardiovascular responses to weight loss interventions [20, 34]. In our analysis, as the credible levels were relaxed, the covariates poor subjective general health and poorly controlled diabetes were consistently the ones to have an expected treatment benefit while the covariates good subjective general health with well-controlled diabetes had an expected null or adverse treatment effect.

The innovation of this paper firstly comes from the inferences made possible by the credible subgroups method: that there is a high posterior probability that all types of patients within the exclusive credible subgroup D experience a positive treatment effect. This addresses a different research question to previous publications that have explored patient heterogeneity but relied on sample average calculations. Our findings compared to those previous demonstrate that it is possible to calculate average estimates that do not capture the responses of minority subgroups with treatment effects that deviate from the mean. Secondly, the credible subgroups method allows us to make statements of simultaneity where all subgroup members experience a treatment effect above some threshold that adjusts for issues of multiplicity due to hypothesis tests in multiple subgroups. This helps to avoid the perception of data dredging which underpins prevalent scepticism of standard subgroup analyses.

Previous research has used exploratory endpoints and *ad hoc* subgroup analysis to identify significant treatment effects reported in secondary studies. Using the original CVD endpoint, our analysis shows that as the credible level is relaxed, the range of the covariate profiles for exclusive subgroup membership expands. There exists a trade-off between statistical confidence and subgroups too narrowly defined to represent any application in medical care. As the credible level is relaxed from 80% to 50%, the proportion of covariate points classified to the exclusive credible subgroup increases. Our research identifies a minority subgroup consisting of covariate profiles with an expected treatment benefit. Based on an 80% credible level, these covariate profiles are used to classify less than 0.5% of the Look AHEAD trial who would simultaneously be expected to experience a positive treatment effect. This contradicts previous research stating 85% of Look AHEAD participants did benefit from treatment as identified by positive average estimates [23].

The findings of this secondary analysis are not meant to overshadow the conclusions from the original Look AHEAD trial. Rather, we demonstrate an alternative method of exploring treatment effect heterogeneity that does not suffer from the limitations of standard subgroup analyses. The credible subgroups method defines subgroups, based on a number of baseline covariates, which consist of all the types of patients expected to benefit or not benefit from a given treatment while accounting for multiplicity. This research applies the credible subgroups method to the Look AHEAD trial to illustrate the trichotomy of the covariate space and identifies a large number of covariate points located in an uncertainty region. This combined with the restrictive covariate profiles of those who would be classified to the exclusive credible subgroup, explains how so few patients with an expected treatment benefit could have been lost in trial-wide average estimates.

This analysis of the Look AHEAD trial has some limitations. At first glance the output from the credible subgroups method may not be as straightforward to interpret compared to standard subgroup analyses, but the fundamental objective of identifying individual covariate profiles of patients expected to benefit cannot be achieved through simple average calculations. Secondly, the credible subgroups method is not designed for variable selection. Instead, the credible subgroups method requires a list of covariates believed to have predictive value when interacted with treatment which requires *a priori* knowledge of intervening factors. Thirdly, the credible subgroups method computationally struggles with increasing covariates due to a reduction in statistical power and the inclusion of many continuous covariates [6]. The possible biases produced by these limitations are similar to those found in standard subgroup analyses. Regarding the biological plausibility of the chosen covariates in particular, all are standard medical and demographic variables in studies examining behavioural treatments for type 2 diabetes and the confounding effects of obesity [14, 17, 18, 20, 23, 28]. We conclude that covariates previously identified as being important for exploring treatment heterogeneity may only explain a small portion of the entire covariate space that is useful for identifying positive treatment effects. In fact, variable selection itself introduces multiplicity problems, and accounting for this source of multiplicity as well would necessitate even more cautious conclusions. In particular, the inconclusive region (D \ S) would need to be enlarged for any given credible level. Some study of variable selection via Stochastic Search Variable Selection (SSVS) within the regression step of the credible subgroups approach has been done [35], which shows this effect in the sense that D \ S is larger when not conditioning on a selected model. As this relates to the comparison with Baum et.al. [23], we have shown that accounting for only some but not all sources of multiplicity does not allow as optimistic conclusions as the previous analysis. This result would be made even more extreme by accounting for additional sources of multiplicity. Finally, point estimates for treatment effects in the selected subgroups D and S-complement are biased away from zero due to selection bias. Thus conclusions drawn from the credible subgroups method should be limited to hypothesis testing, and plots such as Figure 3 should be interpreted only as diagnostics and not as post-selection effect estimates.

There exists much scope for future research in the synergy of the credible subgroups method into adaptive enrichment trial design. The objectives of adaptive trial design and the credible subgroups method are the same: to increase the likelihood of identifying a true subgroup of patients expected to benefit from a given therapy [36]. Adaptive trial design includes prospectively planned analyses that can change the course of an on-going trial based on accumulating the data of subjects within the trial [37]. Adaptive trial design stands to benefit from the credible subgroups method if used throughout interim analyses and at early stage enrolee selection. Sequential trial design permits stopping early for efficacy based on interim analyses [38, 39], which could be defined by the trichotomy of the covariate space using the credible subgroups method. Overwhelming evidence of treatment benefit or harm would allow for early termination while the trial would continue to gain information about the covariate space where classifications are uncertain. Additionally, the credible subgroups method could be used in an adaptive population-enrichment design where biomarkers are identified that would predict response or lack of response to a given therapy [40]. This information could be used in confirmatory trials to limit entry of patients to stage 3 randomisation [41]. In this research, biomarkers for diabetes and obesity that are related to the effectiveness of the lifestyle intervention would be of particular interest. Future incorporation of the credible subgroups method into adaptive enrichment trail design can magnify the potential treatment effect, reduce costs and increase trial efficiency.

## Data Availability

Legal requirements prohibit public sharing of the dataset. Data are available upon request from the National Institute of Diabetes and Digestive and Kidney Diseases (NIDDK). AC and JF had full
access to all of the data and the final responsibility to submit for publication.

## 6. Acknowledgements

The work of AC and JF was funded by the Health Research Board (grant number HRB-RL-2013-11). The work of JS was funded by the National Institute for Health Research Biomedical Research Centre at the University of Oxford (grant number NIHR-BRC-1215-20008).

Data are available upon request from the National Institute of Diabetes and Digestive and Kidney Diseases (NIDDK) at https://www.niddk.nih.gov/.

## 6. Supplement

**S_Fig1. Trichotomy of Baseline Covariates at 80% Credible Level with Vague Prior on Predictive and Prognostic Effects**.

**S_Fig2. Trichotomy of Baseline Covariates at 65% Credible Level**

